# Application of the Health Belief Model Using the PRECEDE–PROCEED Framework for Community-Based Hypertension Control: A Cluster Randomized Trial

**DOI:** 10.64898/2026.06.23.26356314

**Authors:** Mohammad Tanvir Islam, Md Atiqul Haque, Muhammad Kamal Uddin, Mohan Chakraborty, Shohael Mahmud Arafat

## Abstract

**Background:** Hypertension is a leading cause of cardiovascular morbidity and premature death in low- and middle-income countries (LMICs), where blood-pressure control remains poor because of suboptimal medication adherence, unhealthy lifestyle behaviours, and structural barriers to care. Theory-driven interventions based on the Health Belief Model (HBM) have shown promise, but most have treated the six HBM constructs as interchangeable predictors. Formative mixed-methods work by our group in the same population indicates that the constructs do not operate uniformly: perceived barriers and cues to action are associated with systolic blood pressure (SBP) through medication adherence, self-efficacy is associated with SBP through pathways other than adherence, and perceived severity—though near-universally endorsed—is motivationally inert. This trial will evaluate an HBM-based, mediation-informed, PRECEDE–PROCEED-guided educational intervention prioritised according to that mechanistic evidence.

**Methods:** This is a parallel-group, community-based cluster randomised controlled trial with 1:1 allocation. Twelve community clusters in Kamalganj sub-district, north-east Bangladesh, will be randomly assigned to an HBM-based educational intervention or usual care (approximately 480 participants, ∼40 per cluster). The intervention, delivered over 12 months by trained community health workers, comprises five components: (i) group education with weighted emphasis on perceived barriers and self-efficacy; (ii) individual barrier-mapping and motivational counselling; (iii) enabling strategies including community blood-pressure monitoring corners, drug-supply continuity advocacy, and culturally compatible substitution for traditional remedies; (iv) reinforcing strategies through structured family engagement; and (v) provider training on gender-perception bias. The primary outcomes are change in mean SBP and the proportion achieving controlled blood pressure at 12 months. Secondary outcomes include HBM construct scores, medication adherence (Bangladesh Medication Adherence Scale, BMAS), lifestyle behaviours, and use of culturally embedded folk remedies, assessed at baseline, 6 and 12 months. Analyses follow intention-to-treat using mixed-effects models that account for clustering. An exploratory longitudinal mediation analysis will test whether trial-induced change in adherence mediates trial-induced change in SBP.

**Discussion:** This trial will provide a prospective test of a mediation-prioritised HBM intervention and, if effective, a culturally tailored, scalable model for hypertension control in Bangladesh and comparable low-resource settings.

**Trial registration:** This protocol is registered in ClinicalTrials.gov. Identifier: NCT07426978 (Date: 16/02/2026), Unique Protocol ID- PR-1440 (https://register.clinicaltrials.gov/prs/beta/records)

## Introduction

Hypertension is a leading modifiable risk factor for cardiovascular disease, stroke, chronic kidney disease, and premature mortality worldwide [1, 2]. The burden is disproportionately high in low- and middle-income countries (LMICs), where most people with hypertension live and where awareness, treatment, and control rates remain low [3, 4]. In South Asia, rapid urbanisation, population ageing, and shifts in diet and physical activity have driven a steep rise in prevalence, yet large proportions of treated patients have uncontrolled blood pressure [5].

National surveys in Bangladesh indicate that approximately one in four adults has hypertension and that fewer than half of those treated achieve blood-pressure control [5, 6]. Barriers operate at individual, health-system, and contextual levels, including poor medication adherence, high dietary salt intake, low fruit consumption, physical inactivity, tobacco use, irregular follow-up, fragmented care pathways, and limited availability of affordable antihypertensive medicines [7–10]. These behavioural and environmental determinants are strongly shaped by patients’ health beliefs, perceived barriers, and self-efficacy, yet routine hypertension care in Bangladesh rarely addresses these psychosocial factors in a structured way.

Theory-informed interventions have been advocated to promote sustained behaviour change in chronic-disease management [11]. The Health Belief Model (HBM) is among the most widely used frameworks for explaining and modifying preventive and treatment behaviours, positing that action is influenced by perceived susceptibility, perceived severity, perceived benefits, perceived barriers, cues to action, and self-efficacy [12]. HBM-based educational programmes have improved medication adherence, lifestyle modification, and blood-pressure control among people with hypertension, including in LMIC settings [13]. The PRECEDE–PROCEED model complements the HBM by providing a structured framework for the participatory assessment of social, epidemiological, behavioural, and environmental determinants (PRECEDE) and for guiding implementation and multi-level evaluation (PROCEED) [14, 15].

Our formative mixed-methods study, conducted in Kamalganj sub-district among 461 hypertensive adults, provided mechanistically detailed insight into how the HBM operates in this population (Islam et al., preprint; DOI forthcoming) [16]. Three findings directly inform the present protocol. First, perceived barriers and cues to action were associated with SBP indirectly through medication adherence, with perceived barriers emerging as the single strongest correlate of optimal adherence; barrier reduction is therefore the most powerful modifiable lever in this setting. Second, self-efficacy was associated with lower SBP through pathways other than medication adherence, suggesting that confident self-managers achieve control through multi-domain self-care—diet, blood-pressure self-monitoring, stress regulation, and physical activity—rather than pill-taking alone. Third, perceived severity, although near-universally endorsed, did not predict adherence and showed an inverse adjusted association, a fear-without-efficacy pattern consistent with Witte’s Extended Parallel Process Model [17]; perceived benefits predicted adherence but not SBP, plausibly because culturally embedded practices (lemon-water with salt and sugar, roasted salt, oral saline as a beverage) and treatment-quality issues dilute the clinical translation of belief-driven adherence.

Taken together, these findings reframe the HBM as a multi-channel rather than a single belief-to-outcome pipeline and argue for a strategic re-prioritisation of intervention effort: barrier reduction first, domain-specific self-efficacy second, interpersonal cues third, with severity-based fear messaging replaced by enablement and benefit messaging complemented by explicit cultural-practice substitution. Few interventions to date have integrated such within-population mechanistic evidence with the structured planning of PRECEDE–PROCEED. This trial therefore aims to evaluate a mediation-informed, HBM-based, PRECEDE–PROCEED-guided educational intervention for improving medication adherence and blood-pressure control among community-dwelling hypertensive adults.

### Objectives

#### General objective

To evaluate the efficacy of a mediation-informed, Health Belief Model-based educational intervention, developed using the PRECEDE–PROCEED framework, in improving blood-pressure control and medication adherence among community-dwelling adults with hypertension.

#### Specific objectives

- To determine the effect of the intervention on mean systolic and diastolic blood pressure at 6 and 12 months.
- To compare the proportion of participants achieving controlled blood pressure between arms at 6 and 12 months.
- To assess changes in HBM construct scores—particularly perceived barriers and self-efficacy as the empirically prioritised constructs—from baseline to 6 and 12 months.
- To evaluate the effect of the intervention on medication adherence (BMAS) over 6 and 12 months.
- To evaluate changes in lifestyle behaviours, including sodium intake, physical activity, fruit and vegetable consumption, and tobacco use.
- To measure change in self-reported use of culturally embedded folk remedies (lemon-water, roasted salt, oral saline).
- To explore whether trial-induced change in medication adherence mediates trial-induced change in SBP, replicating longitudinally the cross-sectional mediation pattern observed in formative work.
- To conduct a process evaluation of intervention fidelity, reach, dose delivered, dose received, and acceptability.

## Methods

This protocol is reported in accordance with the Standard Protocol Items: Recommendations for Interventional Trials (SPIRIT) 2013 statement; a completed SPIRIT checklist is provided as a supporting file, and the schedule of enrolment, interventions, and assessments is summarised in Fig 1 (SPIRIT figure). The trial will be reported in line with the CONSORT 2010 extension for cluster randomised trials.

**Fig 1.** Schedule of enrolment, interventions, and assessments (SPIRIT figure). The figure summarises the study periods—pre-trial PRECEDE assessment and intervention finalisation, enrolment and cluster allocation, the 12-month intervention, and outcome assessment at baseline, 6 months, and 12 months—and the timing of each measurement (blood pressure, HBM construct scores, BMAS adherence, lifestyle behaviours, cultural-practice indicators, and process-evaluation measures), in accordance with the PRECEDE–PROCEED framework. [Figure file to be supplied at submission.]

### Study design

This is a parallel-group, community-based cluster randomised controlled trial evaluating an HBM-based educational intervention for hypertension, planned using the PRECEDE–PROCEED framework and informed by mediation analysis of formative mixed-methods data. Clusters (villages or wards) will be allocated 1:1 to intervention or control, with outcome assessment at baseline, 6 months, and 12 months. The cluster design minimises contamination between participants and accommodates community-level components such as group education and community blood-pressure monitoring corners.

### Study setting

The trial will be conducted in Kamalganj sub-district, Moulvibazar district, north-eastern Bangladesh, comprising rural and peri-urban communities served by government community clinics and primary health centres. The area has existing non-communicable-disease surveillance data and was the site of the formative mixed-methods study from which the mediation evidence underpinning this protocol was derived; this continuity strengthens the contextual validity of the intervention design.

### Cluster definition and selection

A cluster is defined as a village or ward with approximately 3,000–5,000 residents and clear geographic boundaries to minimise overlap. Eligible clusters will be identified from administrative records and maps. From the sampling frame, 12 clusters (6 intervention, 6 control) will be purposively selected in collaboration with local health authorities to ensure implementation feasibility and then randomly allocated to study arms.

### Participant eligibility and recruitment

Adults are eligible if they (1) are aged ≥18 years; (2) have physician-diagnosed hypertension or documented systolic BP ≥140 mmHg or diastolic BP ≥90 mmHg on at least two screening occasions; (3) have resided in the selected cluster for ≥6 months and intend to remain during the study period; and (4) are able to provide informed consent. Exclusion criteria are severe cognitive impairment, serious comorbidity requiring intensive care (e.g., advanced cancer, end-stage renal disease), and current pregnancy or intention to become pregnant within 12 months.

Potential participants will be identified from the formative-study baseline register and clinic registers. Community meetings will be held in each cluster, where the study purpose, procedures, risks, and benefits will be explained in Bengali. Interested individuals will be screened for eligibility, and written informed consent (thumb-impression with witness for non-literate participants) will be obtained before enrolment.

### Randomisation, allocation concealment, and blinding

Clusters will be randomly allocated 1:1 to the intervention or control arm using simple randomisation. A computer-generated random sequence will be created by an independent statistician not involved in recruitment or intervention delivery. Allocation codes will be stored in a password-protected file and released only after all clusters are enrolled, so recruiters remain unaware of upcoming assignments. Outcome assessors and data analysts will be blinded to allocation through anonymised study IDs and datasets without group labels, and staff delivering the education will not conduct follow-up measurements. Participants and community health workers cannot be blinded; both arms will receive the same usual hypertension care, with only intervention clusters receiving the additional HBM-based education. If unblinding of blinded personnel becomes essential, the principal investigator will authorise limited access and document the reason, timing, and individuals involved.

### Sample size

The sample size was calculated to detect a between-group difference of 5–7 mmHg in mean systolic BP at 12 months, assuming a standard deviation of 20 mmHg, 80% power, a two-sided alpha of 0.05, and an intracluster correlation coefficient (ICC) of 0.02–0.05 based on previous community hypertension trials in similar settings. With an average of 40 participants per cluster, 12 clusters (6 per arm) yield approximately 480 participants and provide adequate power for the primary BP outcomes and moderate effect sizes for secondary outcomes such as HBM construct scores and adherence. The sample is also adequate for exploratory longitudinal mediation analyses (≥10 events per parameter). Allowance has been made for up to 15% loss to follow-up.

### Theoretical framework and intervention development

Intervention development followed the PRECEDE phases, using completed mixed-methods formative research to identify the social, epidemiological, behavioural, and environmental determinants of poor BP control. Critically, this iteration incorporates evidence from covariate-adjusted mediation analyses of the formative quantitative dataset, which reordered the priority weights assigned to each HBM construct. The mediation findings informed three structural design decisions: (i) perceived barriers and self-efficacy were elevated to the central organising constructs, as they emerged as the strongest empirical levers on SBP; (ii) perceived severity was de-emphasised as a primary target, given its inert and inverse adjusted association with adherence, and is retained only as factual background rather than as a motivational lever; and (iii) perceived-benefits content was deliberately complemented by explicit cultural-practice substitution, given the empirical pattern that benefit beliefs drove adherence but not BP. Behavioural targets identified in formative work—medication non-adherence, high-salt intake, inadequate physical activity, low fruit consumption, tobacco use, and irregular follow-up—and environmental barriers—distance to facilities, work-related time constraints, periodic medicine scarcity at the Upazila Health Complex (UHC), and fragmented care across registered physicians, village doctors, and pharmacy drug-sellers—were mapped onto HBM constructs and onto enabling and reinforcing factors, with mediation-evidence weights reflected in the relative emphasis of each component.

#### PRECEDE Phase 1: Social assessment

Formative work indicated that rural hypertensive individuals perceive themselves as susceptible to hypertension and its cardiovascular consequences and identify mental stress as an important contributor, while their understanding of the preventive role of healthy diet and physical activity is inconsistent. Conventional treatment is valued but its uptake is compromised by cost, distance, and scarcity of free medicines. The health of family members acts as a cue for healthy practices. Qualitative interviews revealed culturally embedded practices (lemon-water, roasted salt, oral saline) coexisting with biomedical treatment that may dilute pharmacological effectiveness.

#### PRECEDE Phase 2: Epidemiological assessment

The community faces a substantial burden of uncontrolled hypertension. In the formative survey, a majority of participants had uncontrolled BP and fewer than half reported optimal medication adherence; most came from lower socioeconomic backgrounds and had minimal formal education, and obesity and diabetes were common. These figures define the epidemiological need addressed by the intervention.

#### PRECEDE Phase 3: Behavioural and environmental assessment

##### Behavioural factors

Beyond medication non-adherence, the formative sample showed inadequate fruit consumption, high-salt behaviour, inadequate physical activity, smoking, and a notably high prevalence of smokeless-tobacco use.

##### Environmental factors

Participants frequently sought care from pharmacy drug-sellers or village doctors without structured counselling while also consulting registered physicians, producing fragmented management. Geographical distance and clinic hours conflicting with work limited regular follow-up, and periodic medicine shortages at public facilities increased reliance on free health camps. These patterns underscore the need for multilevel interventions addressing beliefs, access barriers, and social support.

#### PRECEDE Phase 4: Educational and ecological assessment (HBM integration)

Behavioural and environmental determinants were categorised into predisposing, enabling, and reinforcing factors, with HBM constructs as the guiding framework and the relative emphasis of each construct informed by the empirical mediation hierarchy (Table 1). Predisposing factors comprise the six HBM constructs. Perceived threat (susceptibility and severity) is addressed factually and immediately paired with self-efficacy and barrier-reduction content; fear-based framing is explicitly avoided. Perceived benefits are addressed at moderate priority with an explicit cultural-overlay component. Perceived barriers and self-efficacy are the highest-priority constructs, and cues to action are addressed through locally appropriate interpersonal channels. Enabling factors include domain-specific skills (with pictorial materials, blister-pack reminders, and tactile medication identifiers for low-literacy patients), links to affordable medication, promotion of low-cost seasonal produce, community blood-pressure monitoring corners, and strengthened linkages between community health workers and formal facilities. Reinforcing factors centre on structured family engagement, provider education on gender-perception bias, regular follow-up contacts, and cultural-practice substitution through structured group discussion.

**Table 1.**
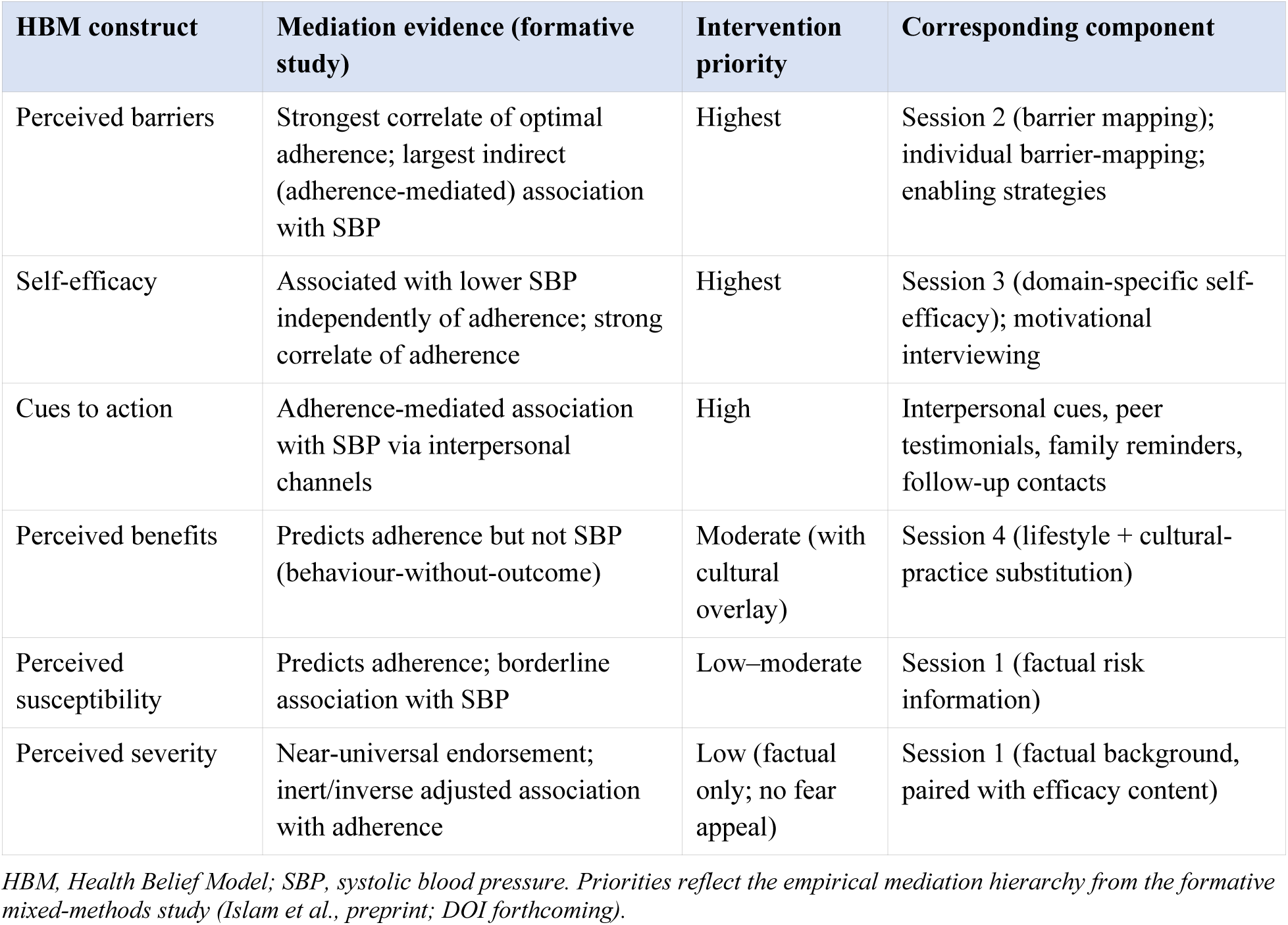
HBM constructs mapped to mediation evidence, intervention priority, and corresponding intervention components.

| HBM construct | Mediation evidence (formative study) | Intervention priority | Corresponding component |
| --- | --- | --- | --- |
| Perceived barriers | Strongest correlate of optimal adherence; largest indirect (adherence-mediated) association with SBP | Highest | Session 2 (barrier mapping); individual barrier-mapping; enabling strategies |
| Self-efficacy | Associated with lower SBP independently of adherence; strong correlate of adherence | Highest | Session 3 (domain-specific self-efficacy); motivational interviewing |
| Cues to action | Adherence-mediated association with SBP via interpersonal channels | High | Interpersonal cues, peer testimonials, family reminders, follow-up contacts |
| Perceived benefits | Predicts adherence but not SBP (behaviour-without-outcome) | Moderate (with cultural overlay) | Session 4 (lifestyle + cultural-practice substitution) |
| Perceived susceptibility | Predicts adherence; borderline association with SBP | Low–moderate | Session 1 (factual risk information) |
| Perceived severity | Near-universal endorsement; inert/inverse adjusted association with adherence | Low (factual only; no fear appeal) | Session 1 (factual background, paired with efficacy content) |

*HBM, Health Belief Model; SBP, systolic blood pressure. Priorities reflect the empirical mediation hierarchy from the formative mixed-methods study (Islam et al., preprint; DOI forthcoming).*

#### PRECEDE Phase 5: Administrative and policy assessment

Bangladesh has an extensive network of community clinics staffed by community health-care providers and supported by health assistants and family welfare visitors, with non-communicable-disease (NCD) Corners in health complexes providing free treatment; these cadres can deliver the intervention, supported by local NGOs experienced in health promotion. Resource needs include community health workers and volunteers (with modest incentives), validated BP devices, Bengali educational materials emphasising the prioritised constructs, family-engagement and cultural-practice materials, reminder cards, and mobile phones; an initial five-day training workshop and ongoing supervision; and rigorous expense tracking to inform decisions about wider implementation. Integration with existing community health-worker programmes and reliance on inexpensive, locally sourced materials support sustainability and scalability.

### Intervention and control conditions (PROCEED Phase 6)

The intervention will improve BP control by using HBM constructs as the theoretical framework and by addressing enabling and reinforcing factors through five integrated components, with the relative emphasis of each informed by the empirical mediation hierarchy. It will be delivered over 12 months by trained community health workers. The control arm will receive usual care—standard clinic visits, usual prescribing, and routine counselling—reflecting current standard management; this is ethically appropriate because all participants continue to receive standard care, and it enhances external validity. No restrictions will be placed on routine medical treatment in either arm.

#### Component 1: Group education sessions (mediation-weighted)

Five group sessions will be delivered over a 10-week period (one every two weeks), each lasting 60–75 minutes, at a designated NGO office suitable for lectures and group discussion. The curriculum is reorganised by mediation-evidence priority:

- **Session 1 — Understanding hypertension.** What hypertension is, its asymptomatic nature, the importance of regular monitoring and follow-up, and an overview of the multi-channel pathway from beliefs to clinical control. Severity content is delivered factually—not as a fear appeal—and paired immediately with what participants can do.
- **Session 2 — Mapping and overcoming barriers (priority session).** Dedicated to perceived barriers, the strongest empirical lever. Participants identify personal barriers (cost, distance, time, family support, drug supply, fragmented care) and co-develop practical strategies—use of nearby community clinics, convenient scheduling, free-medication enrolment at the NCD Corner, family-mediated medication management, transport pooling—using a barrier-mapping worksheet.
- **Session 3 — Building self-efficacy across multiple domains (priority session).** Dedicated to self-efficacy, the strongest direct contributor to BP control. Each self-management domain—medication, diet, BP monitoring, physical activity—receives domain-specific skill-building, with particular attention to the empirically weakest domains (independent medication recall and regular exercise), including hands-on practice with home BP monitors and context-appropriate exercise.
- **Session 4 — Lifestyle and cultural-practice substitution.** Salt reduction, physical activity, healthy eating, and tobacco cessation, plus explicit content on folk practices that may dilute clinical effectiveness (lemon-water with salt and sugar, roasted salt, oral saline as a beverage) and culturally compatible substitutes.
- **Session 5 — Family engagement.** A dedicated session for family members (spouses, adult children), educating them about hypertension and equipping them with specific skills—medication recognition by colour or shape, dosing schedules, BP monitoring, and warning symptoms requiring care-seeking.

Sessions will use interactive lectures with visual aids, group discussion, role-play, barrier-mapping worksheets, and self-efficacy skill demonstrations, integrating brief patient testimonials (the empirically strong cue of peer observation) where feasible. Trained community health workers or nurses will facilitate using standardised manuals and checklists, with prior training on the mediation-informed model, motivational interviewing, gender-bias awareness, and cultural-practice substitution; competency will be assessed before implementation and reassessed mid-intervention.

#### Component 2: Individual counselling and motivational interviewing

Three one-on-one sessions at weeks 4, 8, and 16 (each 30–40 minutes) will deliver personalised barrier mapping using the Session-2 worksheet, motivational-interviewing techniques to strengthen self-efficacy in the weakest domains, explicit screening for and discussion of cultural practices that may neutralise adherence, and goal-setting with a written agreement signed by participant and counsellor. Trained community health workers will conduct the sessions under ongoing supervision.

#### Component 3: Enabling strategies (resources and skills)

- Community BP monitoring corners equipped with validated automated devices and operated by trained volunteers, where participants check BP monthly, record results, and learn when to seek care.
- Educational materials including printed reminder calendars with medication tick-boxes, pictorial Bengali leaflets on low-salt recipes and culturally compatible substitutes, and wallet cards summarising key messages.
- Mobile-phone reminders (where access permits) as a supplement to—not a replacement for—interpersonal cues, given the limited reach of digital channels in this population.
- Linkages to affordable medication, including information on government essential-drug programmes and collaboration with local providers to encourage monitoring and counselling by trained personnel.
- System-level advocacy for stable NCD Corner drug supply, with the team monitoring and reporting drug-supply continuity to address periodic stockouts identified as a critical structural barrier.

#### Component 4: Reinforcing strategies (social support and feedback)

- Family involvement through at least two dedicated family-engagement sessions (Session 5 and a month-6 refresher), with materials enabling family members to support adherence, low-salt cooking, and BP monitoring.
- Follow-up contacts: monthly phone calls or home visits by community health workers during months 3–12 to encourage, troubleshoot, assess progress, and reinforce key messages.
- Peer support, where feasible, through informal networks or walking groups, leveraging the empirically strong cue of peer observation.
- Recognition of participants who achieve target BP or demonstrate sustained change, through certificates or public recognition at community events.

#### Component 5: Provider training on gender-perception bias

A two-day provider workshop for community health workers and pharmacy drug-sellers in intervention clusters at study commencement, with a half-day refresher at month 6, will address the empirically documented gender-perception bias whereby providers characterise women as more negligent despite quantitative evidence to the contrary. Content includes recognition of the bias, role-plays to develop gender-equitable counselling, and case discussions.

### Outcomes and measurement (PROCEED Phase 7)

#### Primary outcomes

- Change in mean systolic blood pressure from baseline to 12 months.
- Proportion of participants with controlled blood pressure (systolic <140 mmHg and diastolic <90 mmHg) at 12 months.

#### Secondary outcomes

- Change in mean diastolic blood pressure from baseline to 12 months.
- Blood-pressure control at 6 months (intermediate outcome).
- Change in BMAS adherence score and adherence category.
- Change in HBM construct scores, particularly perceived barriers and self-efficacy.
- Change in lifestyle-behaviour scores (salt, physical activity, fruit and vegetable intake, tobacco use).
- Reduction in self-reported use of cultural folk remedies (lemon-water, roasted salt, oral saline).

#### Instruments and procedures

HBM construct scores will be assessed with the validated 38-item HBM questionnaire developed in formative work (Cronbach’s α 0.63–0.77 across most constructs; perceived-severity α reflects a ceiling effect and is supplemented by item-level analysis), scored on a 3-point scale (Agree / Not sure / Disagree). Medication adherence will be measured with the Bangladesh Medication Adherence Scale (BMAS), a validated 9-item, three-factor instrument (Behavioural, Economic, Cognitive; total 0–27) [18], with optimal/sub-optimal classification and self-reported missed doses over the past month. Lifestyle behaviours, cultural-practice indicators, health-service utilisation, and hypertension knowledge will be captured with structured items administered at baseline (before randomisation), 6 months, and 12 months by trained interviewers blinded to allocation where feasible.

##### Blood-pressure measurement

Blood pressure will be measured by trained data collectors using validated automated oscillometric devices (OMRON HEM-7120). Participants will rest seated for at least five minutes; two readings will be taken on the same arm 1–2 minutes apart and averaged. Measurements will follow standardised WHO/ISH procedures and devices will be calibrated regularly [19].

### Process evaluation

Following the PROCEED framework, the process evaluation will assess fidelity (proportion of planned sessions delivered and adhering to the curriculum, with specific attention to barrier-mapping and self-efficacy components), reach (proportion of eligible individuals enrolled and attending ≥75% of sessions, representativeness, and family-session coverage), dose (sessions attended, counselling and follow-up contacts received, BP checks, and exposure to cultural-practice content), acceptability (post-intervention interviews or focus groups with participants, family members, and staff), provider gender-bias outcomes (pre/post-training assessment), and contamination (exit interviews and field reports). Data sources include attendance registers, facilitator checklists, supervisor observation forms, participant questionnaires, and field notes.

### Data management and statistical analysis

Data will be double-entered using REDCap with validation rules, regular quality checks, secure password-protected storage, and cleaning before analysis. Primary analyses will follow the intention-to-treat principle, including all randomised participants in their allocated clusters, using mixed-effects regression models that account for clustering. Continuous outcomes (SBP, adherence score, HBM construct scores) will be analysed with linear mixed-effects models with random cluster intercepts and adjustment for baseline values; binary outcomes (BP control, optimal adherence, cultural-practice prevalence) with generalised linear mixed-effects models with a logit link. Missing data will be handled primarily through these models, which use all available observations; if missingness is substantial, multiple imputation and complete-case analyses will be performed as sensitivity checks. Pre-specified exploratory analyses include per-protocol analyses and subgroup analyses (by sex, age, and socioeconomic status) using interaction terms.

#### Exploratory mediation evaluation

Longitudinal mediation analyses will test whether trial-induced change in the BMAS adherence score mediates trial-induced change in mean SBP at 12 months, using a parallel-process latent growth model and a counterfactual-based mediation framework [20, 21] to estimate the average causal effect of intervention assignment on adherence change, the effect of adherence change on SBP change, and the indirect and direct effects of assignment on SBP change. This longitudinal replication of the cross-sectional mediation finding would, to our knowledge, constitute the first prospective demonstration of the multi-channel HBM in a hypertension intervention trial. Sensitivity analyses will examine departures from key assumptions (sequential ignorability, stable-unit-treatment-value).

### Harms, monitoring, and trial oversight

The health-education intervention is minimal risk. Any unintended effects reported by participants will be documented and managed, with referral to health facilities as needed and prompt reporting of serious adverse events to the ethics committee. Given the minimal-risk, educational nature of the trial, no formal independent Data Monitoring Committee will be established; oversight will be provided by the principal investigator and a small steering group without relevant financial conflicts of interest. No formal interim efficacy analyses or statistical stopping rules are planned; early termination would be considered only by the principal investigator in consultation with the ethics committee if unexpected safety or feasibility issues arise. Trial conduct and safety will be reviewed internally every 3–6 months using recruitment logs, attendance records, adverse-event reports, and data-quality checks; no external monitoring visits are planned.

### Study timeline and status

The proposed start date is 1 July 2026 and the estimated end date is 30 June 2028 (24-month total duration). The study will proceed through a pre-trial PRECEDE assessment and intervention finalisation, enrolment and cluster allocation, 12 months of intervention delivery, and outcome assessment at baseline, 6 months, and 12 months. The study has not yet started; this document presents the trial protocol revised in accordance with the formative mixed-methods study findings.

### Ethics and dissemination

The protocol and consent materials will be approved by the Institutional Review Board (IRB) of Bangladesh Medical University before recruitment, and any major amendments will be re-approved and communicated to study staff, the trial registry, and participants as needed. The trial will be conducted in accordance with the Declaration of Helsinki and national regulations. Trained research staff will obtain written informed consent (thumb-impression with witness for non-literate participants). Identifiable information will be stored separately from coded study data in locked cabinets and password-protected, access-restricted databases, and only aggregate, non-identifiable results will be reported. No ancillary sub-studies or biological specimens are planned. Participants will continue to receive usual hypertension care, and any harm related to trial procedures will be managed and compensated according to institutional and national policies. Findings will be disseminated through peer-reviewed publications, conference presentations, and summary briefs for local health authorities, providers, and communities.

## Data Availability

Data sharing is not applicable to this protocol, as no datasets were generated or analysed. On completion of the trial, de-identified data, the data dictionary, the statistical analysis plan, and study materials will be available from the corresponding author on reasonable request, subject to Bangladesh Medical University IRB approval and a signed data-access agreement, for non-commercial research.

## Declarations

## Ethics approval and consent to participate

Ethical approval will be obtained from the Institutional Review Board of Bangladesh Medical University (BMU; formerly Bangabandhu Sheikh Mujib Medical University) before recruitment. The trial will be conducted in accordance with the Declaration of Helsinki. Written informed consent (or witnessed thumb-impression where literacy is limited) will be obtained from all participants.

## Trial registration

The protocol will be registered at ClinicalTrials.gov after IRB approval; the registration number will be added on registration.

## Consent for publication

Not applicable; no individually identifiable data are reported in this protocol.

## Author contributions

**Conceptualization:** Mohammad Tanvir Islam, Shohael Mahmud Arafat. **Methodology:** Mohammad Tanvir Islam, Muhammad Kamal Uddin, Md Atiqul Haque. **Formal analysis:** Mohammad Tanvir Islam, Muhammad Kamal Uddin. **Investigation:** Mohammad Tanvir Islam, Md Atiqul Haque, Mohan Chakraborty. **Writing – original draft:** Mohammad Tanvir Islam. **Writing – review & editing:** Md Atiqul Haque, Muhammad Kamal Uddin, Mohan Chakraborty, Shohael Mahmud Arafat. **Supervision:** Shohael Mahmud Arafat. All authors read and approved the final manuscript.

## Acknowledgements

The authors thank the community health workers, local health authorities, and community members of Kamalganj sub-district for their support of this study.

